# Mid-life association between cardiovascular risk factors and cerebral blood flow in a multi-ethnic population

**DOI:** 10.1101/2024.10.04.24314929

**Authors:** Esther M.C. Vriend, Mathijs B.J. Dijsselhof, Thomas A. Bouwmeester, Oscar H. Franco, Henrike Galenkamp, Didier Collard, Aart J. Nederveen, Bert-Jan H. van den Born, Henk J.M.M. Mutsaerts

## Abstract

**Background:** Cardiovascular (CV) risk factors are associated with cerebrovascular damage and cognitive decline in late life. However, it is unknown how different ethnic CV risk profiles are related to cerebral haemodynamics in mid-life. We aimed to investigate associations of CV risk factors with cerebral haemodynamics at two timepoints and examine the impact of ethnicity on these measures.

**Methods:** From the HELIUS study (53.0 years, 44.8% female), participants of Dutch (n=236), Moroccan (n=122), or South-Asian Surinamese (n=173) descent were included. Cerebral blood flow (CBF) and its spatial coefficient of variation (sCoV, marker of macrovascular efficiency) were obtained in both grey (GM) and white matter (WM). Associations of CV risk factors, WM hyperintensities (WMH), and carotid plaques with cerebral haemodynamics were investigated using linear regressions.

**Results:** CBF and sCoV differed per ethnicity. Only at the second visit associations were found, without an interaction with ethnicity; history of CV disease with lower GM CBF and higher WM sCoV, higher total cholesterol and lower WMH volume with lower WM CBF, smoking with higher WM sCoV, and higher SBP with lower GM sCoV.

**Conclusions:** These findings emphasise the need to further explore the longitudinal effects of midlife risk factors and cerebrovascular health, and its interaction with ethnicity.

## Introduction

Systemic cardiovascular (CV) risk factors exhibit a robust association with cardiovascular disease (CVD), cerebrovascular pathology, and cognitive impairment (1, 2). Unlike white matter hyperintensities (WMH) - a commonly used but relatively late-stage structural indicator of cerebrovascular damage (3) – cerebral haemodynamics may be able to detect pathological cerebral changes earlier (4, 5). Cerebral haemodynamics can be non-invasively assessed using arterial spin labelling (ASL) magnetic resonance imaging (MRI) (6), measuring cerebral blood flow (CBF) and its spatial coefficient of variation (sCoV) as a proxy of macrovascular efficiency (7).

Previous studies investigating the association between CV risk factors and CBF in late-life (>65 years) have shown conflicting results. For example, some studies found a negative longitudinal association between blood pressure (BP) and CBF (8-10), differential relations of systolic BP (SBP), or longitudinal development of SBP with grey matter (GM) CBF (11). Other studies found increases in GM CBF after hypertensive treatment (12), or did not find any associations of BP with GM CBF and sCoV at all (13).

Perhaps, the variability of accumulated cerebrovascular pathology is high in late-life, possibly due to differences in CV risk factor exposure or treatment, and might develop non-linear in time (14). Furthermore, ethnic differences in CV risk factors and CV disease may also impact the association between CV risk factors and CBF (15). This encouraged us to further explore the relationship between mid-life CV risk factors and cerebral haemodynamics over time in a multi-ethnic population.

Hence, within a longitudinal multi-ethnic cohort study, we investigate 1) demographical and ethnic differences in CBF and sCoV, 2) the associations of CV risk factors, carotid plaque presence, and WMH with CBF and sCoV and 3) the impact of ethnicity on these associations.

## Methods

### Study population

Data were derived from the HEalthy Life In an Urban Setting (HELIUS) study. The study design and procedures of the HELIUS study have been described in detail elsewhere (16). Briefly, participants were randomly invited, stratified by ethnic background, using the municipality register of Amsterdam, The Netherlands. Data collection consisted of a questionnaire/interview, physical examination at a research location, and the collection of biological samples. For the present study, conducted in accordance with the STrengthening the Reporting of OBservational studies in Epidemiology (STROBE) guidelines, we used the CV risk factor data from the baseline HELIUS data collection (2011-2015), and the second visit (2019-2022) (Figure 1). Data were collected at both time points using the same methods and procedures.

**Figure 1:**
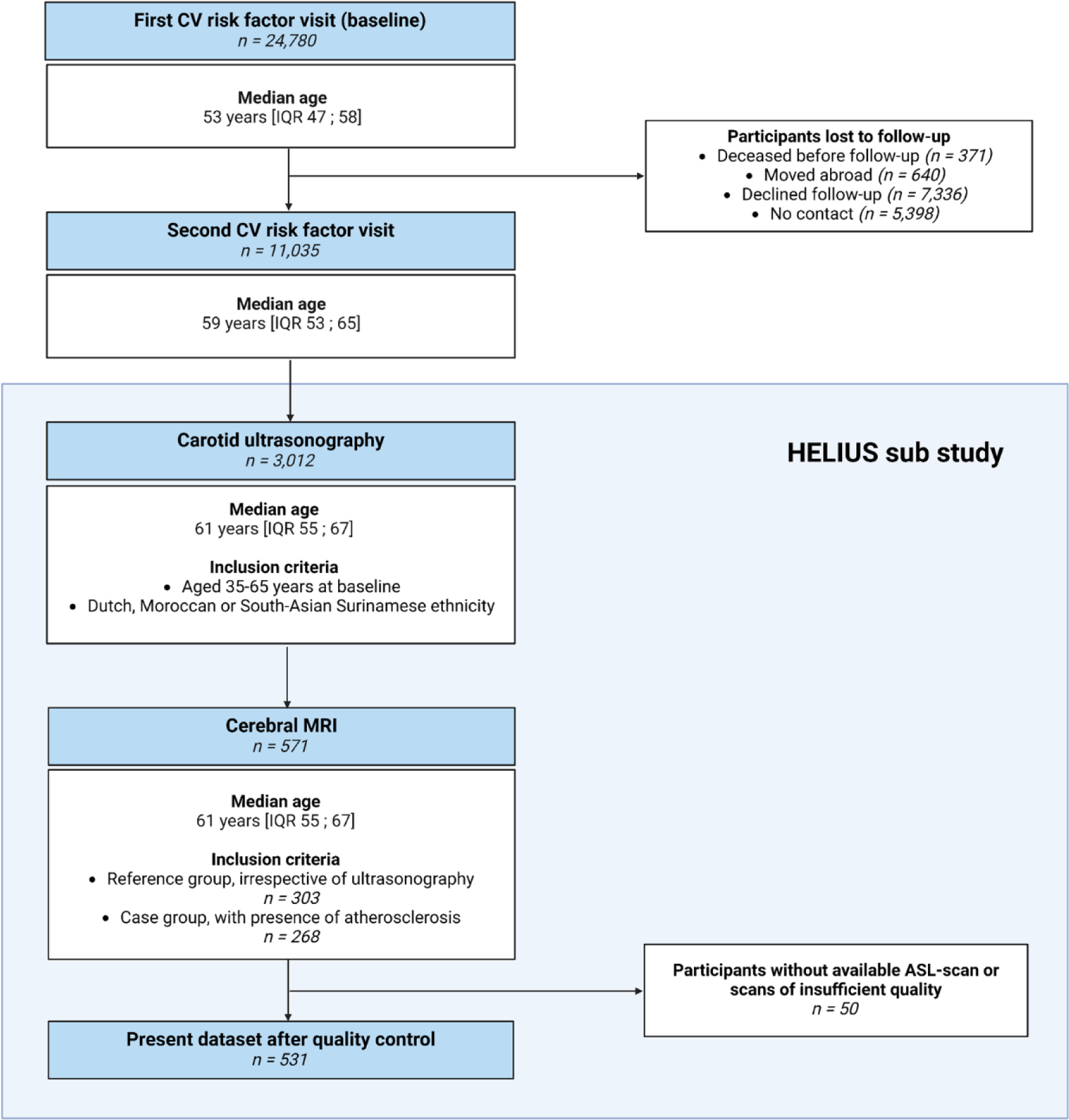
Flowchart of included participants. ASL = arterial spin labelling. HELIUS = Healthy Life in an Urban Setting. IQR = interquartile range. MRI = magnetic resonance imaging.

Between 2021 and 2023, a subset of all HELIUS participants underwent carotid ultrasonography, in which we determined the carotid intima-media thickness (cIMT) and the presence of carotid plaques. Details on the ultrasonography measurements are available elsewhere (17). Inclusion for this substudy was limited to participants aged between 35 and 65 at baseline of Dutch, Moroccan, and South-Asian Surinamese descent with complete baseline and follow-up measurements. A subset of all participants in the carotid ultrasonography substudy received an additional MRI examination (n = 571), from which we used the anatomical and perfusion MRI data. A number of 268 participants were enrolled for MRI examination based on the presence of carotid plaque formation on carotid ultrasound (defined as a carotid plaque of ≥ 2.5 mm on one or both sides or maximum cIMT of >1.0 mm). An additional 303 participants were randomly selected from the carotid ultrasonography sub-study, stratified by ethnic group, as the reference group. Exclusion criteria for MRI comprised unwillingness to participate in the MRI examination or MRI contra-indications.

Due to MRI scanning time constraints, ASL scans were available in 556 individuals. After excluding those with insufficient data quality (arterial transit artefacts and motion artefacts), 531 participants were included in the present study. The MRI examinations were performed at a median of 8.4 years [IQR 7.4 – 9.5 years] after the first CV risk factor visit (baseline) and a median of 2.2 years [IQR 1.8 – 2.6 years] after the second CV risk factor visit. The HELIUS study aligns with the Declaration of Helsinki, and was approved by the Amsterdam UMC, location AMC institutional review board. Written informed consent was obtained from all participants.

### Definitions and measurements

Ethnic background was determined based on the participant’s country of birth and that of their parents. Non-Dutch ethnicity was defined as a participant being born outside the Netherlands and having at least one parent born outside the Netherlands, or if a participant was born in the Netherlands but both parents were born outside the Netherlands. For those of Surinamese ethnicity, further categorization was performed based on their self-reported ethnic origin (‘South-Asian’, ‘African’, or ‘other’). Cardiovascular disease history was defined as a self-reported history of stroke, myocardial infarction, or coronary or peripheral revascularization. Self-reported smoking status was categorized as current, former, or never smokers. Fasting plasma samples were used to measure the concentrations of haematocrit, creatinine, total cholesterol, low-density lipoprotein (LDL) and high-density lipoprotein cholesterol (HDL), and glucose levels. The estimated glomerular filtration rate (eGFR) was calculated using the revised CKD-EPI 2021 creatinine equation (18). Body mass index (BMI) was calculated by dividing measured weight (in kg) by measured height (in m^2^). BP was measured twice on the left arm of participants while seated, after a minimum of 5 minutes of rest, using a validated semi-automatic oscillometric device (Microlife WatchBP Home; Microlife AG, Switzerland).

The average of the two measurements was used to determine SBP and diastolic BP (DBP) levels. Pulse pressure (PP) was defined as the difference between SBP and DBP levels, and mean arterial pressure (MAP) was defined as the DBP plus one-third of the PP. Blood pressure-lowering medication was classified using the Anatomical Therapeutic Chemical classification system. Hypertension was defined as elevated BP levels (≥ 140/90 mmHg) or the use of BP-lowering medication. Diabetes mellitus was defined as elevated fasting glucose levels (≥ 7 mmol/L) and/or the use of glucose-lowering medication.

### MRI acquisition

MRI was performed using a 3T Ingenia scanner (Philips Healthcare, Best, The Netherlands) equipped with a 32-channel head coil. 3D T1-weighted (T1w) magnetization-prepared rapid gradient echo (MP-RAGE) scans were acquired with the following parameters: voxel size = 1x1x1 mm, echo time (TE) = 3.30 ms, repetition time (TR) = 7000 ms, flip angle (FA) = 9 degrees, and inversion time (TI) = 900 ms. 3D fluid-attenuated inversion recovery (FLAIR) scans were obtained with the following parameters: voxel size = 1.10 x 1.10 x 1.12 mm^3^, TE = 356 ms, TR = 4800 ms, FA = 40 degrees and TI = 1650 ms. Pseudo-continuous ASL 2D Echo-Planar Imaging scans were acquired with the following parameters: voxelsize = 3 x 3 x 7 mm^3^, TR = 4445 ms, TE = 17 ms, FA = 90 degrees, post-labelling delay (PLD) range (considering timing differences between 2D slices) = 1800-2574 ms, labelling duration = 1800 ms, 36 averages, background suppression, accompanied by an equilibrium magnetisation image (M0) without labelling or background suppression (TR = 2000 ms).

The images were analysed using ExploreASL version 1.11.0 (19). Briefly, T1-weighted images were used to segment GM, white matter (WM), and cerebrospinal fluid (CSF) using the Computational Anatomy Toolbox 12 (20), while correcting for white matter hyperintensities (WMH) from FLAIR images using the Lesion Segmentation Toolbox (21). Next, ASL images were registered to the T1w images using rigid-body registration, and the equilibrium magnetisation was calculated voxelwise in the brain tissue using the M0 scan. The recommended single-compartment model was used to quantify CBF (22). All images were non-linearly registered to the Montreal Neurological Institute space. CBF and sCoV values were corrected for haematocrit levels obtained at follow-up. CBF values were partial volume corrected. GM and deep WM regions-of-interest (ROIs) were created by combining existing atlases with individual GM and WM segmentations (partial volume > 0.5 in the ASL resolution), after subtraction of WMH partial volume. To avoid GM signal contamination, the WM ROI was eroded by a 4-voxel-sphere to form a deep WM ROI, hereafter referred to as WM. Mean CBF and sCoV were calculated in the total GM and deep WM.

### Statistical analyses

Sample characteristics of the included population were described as mean (SD), median [IQR] or n (%), stratified by ethnicity. All data were tested for normal distribution using the Shapiro-Wilk test and analysed using the appropriate tests (ANOVA, Kruskal Wallis, Chi-squared test) depending on the distribution of the data. We created line diagrams to illustrate the cross-sectional association between age and both CBF and sCoV, stratified by ethnicity. Separate linear regression models were performed to determine the association of the CV risk factors determined at the first and second visit with the outcome measures (GM and WM CBF and sCoV). Initially, risk factor-specific models containing a single CV risk factor were created and adjusted for age, sex, ethnicity, and follow-up time between risk factor measurement and MRI. We investigated the following risk factors: BMI, smoking, diabetes mellitus, SBP, DBP, total cholesterol, eGFR levels, and history of CVD. In participants without prevalent CVD, we also assessed the effect of SCORE2 (the 10-year risk of cardiovascular disease in Europe (23). Subsequently, multivariate models were built, including BMI, smoking, diabetes mellitus, SBP, DBP, total cholesterol, eGFR levels and history of CVD, similarly adjusted for age, sex, ethnicity, and follow-up time between CV risk factor measurement and MRI. Furthermore, we assessed the associations of carotid plaque presence as determined by carotid ultrasonography and log-transformed WMH as determined by MRI with the outcome variables. We tested for ethnicity interactions in the observed associations by performing subgroup analyses. Inverse probability weighting was applied to address disparities in population characteristics between the reference and case group, with weights based on age, sex, and ethnicity, winsorised at 0.1 and 0.9. Repeating the analyses with a group indicator in the regression analysis (0 = reference group, 1 = case group) or without correction for study design did not affect the results (data not shown). We further tested whether the observed observations were attenuated after correction for motion, which was not the case (data not shown). Moreover, we substituted the SCORE2 with the Framingham risk score (FRS) and BP values with MAP, PP and hypertension and evaluated the effect of changes in risk factors over time, corrected for baseline values. Additionally, we investigated whether the results changed after adjusting BP values for the use of anti-hypertensive medication by increasing SBP levels with 10 mmHg and DBP levels with 5 mmHg (24). As prior research has indicated a correlation between body height and carotid plaque presence, we also explored whether the outcome measures were affected by body height and if this impacted the observed associations (17). P < 0.05 was considered statistically significant. As sensitivity analyses, we adjusted the analyses for multiple corrections using the Benjamini-Hochberg Procedure (FDR). Furthermore, models for GM CBF were repeated with and without partial volume correction and with and without correction for haematocrit. Additionally, we repeated the GM CBF analysis with the exclusion of participants with a positive history of CVD. Rstudio (version 4.3.2) was used for all statistical analyses.

## Results

### Population characteristics

Of the 531 participants, 236 individuals were of Dutch, 173 of South-Asian Surinamese, and 122 were of Moroccan descent (Table 1). The median age of the population at the first CV risk factor visit was 53.0 years [IQR 47.0; 58.0], with 44.8% being female. In both the reference and case groups, the prevalence of a positive history of CVD at the first visit was highest in the South-Asian Surinamese group, while mean BMI levels were highest in the Moroccan group. The percentage of smoking participants was highest in the Dutch population, followed by the South-Asian Surinamese and the Moroccans. At the second CV risk factor visit, similar disparities were found. Mean GM and WM CBF values were 55.8 ± 8.5 mL/100g/min and 13.6 ± 3.2 mL/100g/min respectively (Figure 3). Mean sCoV levels were 42.7 ± 6.0 % and 69.6 ± 22.1% for GM and WM, respectively. No significant differences in characteristics between the participants included in the analyses (n = 531) and all participants (n = 571) were found (Supplementary Table 1).

**Figure 2:**
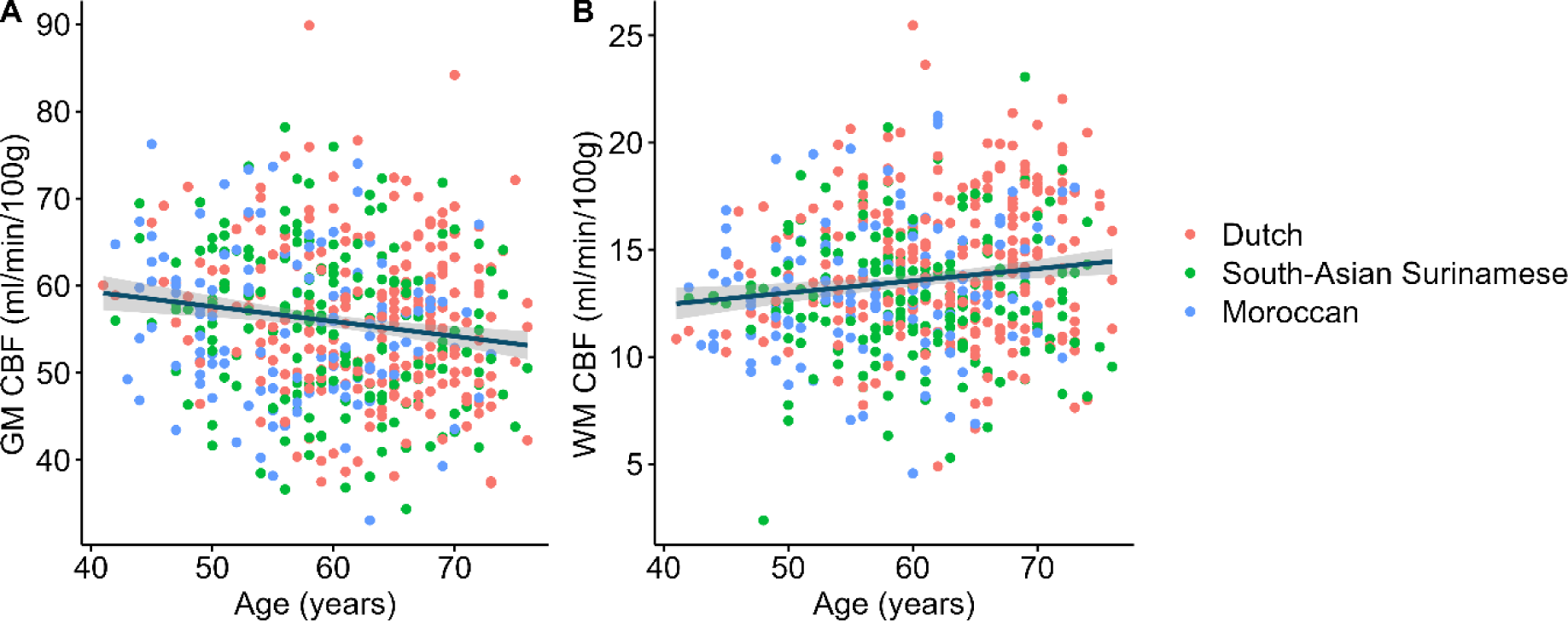
Cross-sectional association between CBF and age in grey matter (A) and white matter (B), stratified by ethnicity. CBF = cerebral blood flow. GM = grey matter. WM = white matter.

**Figure 3:**
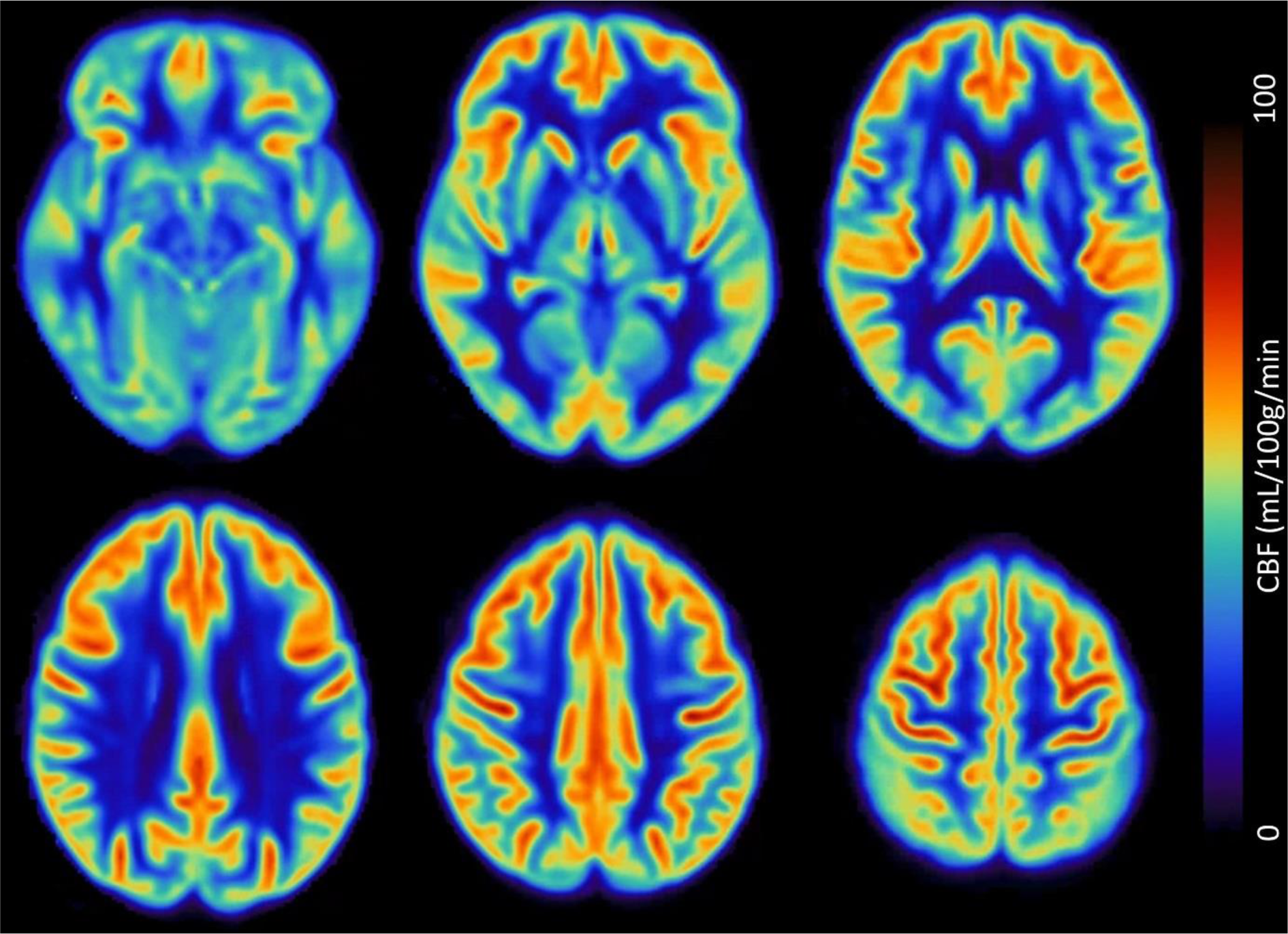
Mean CBF images of participants. Data of n = 531 participants were included. CBF = cerebral blood flow.

**Table 1:** Characteristics of the included population, stratified by ethnic descent. Reference group consists of participants randomly chosen from the HELIUS ultrasonography substudy, stratified by ethnic descent (n = 282). Case groups consist of participants with the presence of atherosclerosis on carotid ultrasound (n = 250). CV = cardiovascular. IQR = interquartile range. SD = standard deviation. BMI = body mass index. SBP = systolic blood pressure. DBP = diastolic blood pressure. CVD = cardiovascular disease. GM = grey matter. WM = white matter. WMH = white matter hyperintensities. CBF = cerebral blood flow. sCoV = spatial coefficient of variation. SA Surinamese = South-Asian Surinamese.

|  | Reference group |  |  | Case group |  |  |
| --- | --- | --- | --- | --- | --- | --- |
|  | Dutch<br>n = 88 | Moroccan<br>n = 98 | SA Surinamese<br>n = 96 | Dutch<br>n = 148 | Moroccan<br>n = 24 | SA Surinamese<br>n = 78 |
| **First CV risk factor visit** |  |  |  |  |  |  |
| Age (years), median [IQR] | 52.0 [45.0, 58.0] | 49.5 [42.0, 55.0] | 50.0 [45.0, 55.0] | 57.0 [52.0, 61.0] | 50.0 [43.8, 53.0] | 53.0 [49.0, 58.8] |
| Sex (female), n (%) | 44 (50.0) | 45 (45.9) | 47 (49.5) | 58 (39.2) | 9 (37.5) | 35 (44.9) |
| SCORE2, mean (SD) | 3.2 (2.1) | 2.6 (1.9) | 3.0 (2.2) | 4.3 (2.3) | 3.4 (1.7) | 3.9 (1.8) |
| BMI (kg/m<sup>2</sup>), mean (SD) | 24.9 (3.8) | 28.5 (4.3) | 25.4 (3.7) | 25.6 (3.6) | 28.3 (4.1) | 25.7 (3.5) |
| SBP (mmHg), mean (SD) | 125.9 (14.1) | 125.6 (15.1) | 129.0 (17.7) | 131.0 (17.8) | 131.3 (15.2) | 137.2 (18.0) |
| DBP (mmHg), mean (SD) | 79.3 (9.7) | 78.4 (7.6) | 82.4 (11.1) | 80.3 (9.7) | 80.8 (7.7) | 84.3 (10.0) |
| Smoking (yes), n (%) | 22 (25.0) | 9 (9.5) | 23 (24.2) | 36 (24.5) | 5 (20.8) | 17 (21.8) |
| Diabetes mellitus, n (%) | 3 (3.4) | 15 (15.5) | 11 (11.8) | 8 (5.4) | 2 (8.3) | 14 (18.2) |
| Tot. cholesterol (mmol/l), mean (SD) | 5.4 (0.9) | 4.8 (0.9) | 5.1 (1.0) | 5.6 (1.0) | 5.2 (1.0) | 5.4 (1.2) |
| History of CVD, n (%) | 1 (1.1) | 2 (2.1) | 9 (9.5) | 8 (5.5) | 0 (0.0) | 9 (11.5) |
| **Second CV risk factor visit** |  |  |  |  |  |  |
| Age (years), median [IQR] | 58.0 [52.0, 64.0] | 55.0 [49.0, 61.0] | 56.0 [50.0, 61.5] | 63.0 [58.0, 67.0] | 55.0 [50.8, 59.0] | 60.5 [55.0, 65.8] |
| SCORE2, mean (SD) | 4.0 (2.3) | 3.8 (2.6) | 4.0 (2.3) | 5.1 (2.3) | 4.3 (2.2) | 5.3 (2.6) |
| BMI (kg/m<sup>2</sup>), mean (SD) | 25.3 (4.1) | 29.1 (4.6) | 26.2 (4.0) | 26.0 (3.6) | 29.0 (4.6) | 25.9 (3.5) |
| SBP (mmHg), mean (SD) | 124.7 (16.1) | 129.1 (16.2) | 126.0 (18.1) | 129.9 (18.0) | 133.4 (16.7) | 134.8 (21.7) |
| DBP (mmHg), mean (SD) | 77.9 (9.5) | 78.1 (10.0) | 79.1 (11.0) | 78.6 (9.3) | 77.4 (9.0) | 80.6 (12.1) |
| Smoking (yes), n (%) | 16 (18.2) | 7 (7.1) | 21 (22.1) | 20 (13.5) | 6 (25.0) | 16 (20.5) |
| Diabetes mellitus, n (%) | 3 (3.4) | 21 (21.4) | 19 (20.0) | 12 (8.2) | 3 (12.5) | 20 (26.0) |
| Tot. cholesterol (mmol/l), mean (SD) | 5.8 (1.2) | 5.1 (1.0) | 5.3 (1.2) | 5.7 (1.2) | 5.3 (1.0) | 5.2 (1.3) |
| Haematocrit (%), mean (SD) | 44.3 (3.0) | 43.3 (4.2) | 43.8 (4.0) | 44.7 (3.1) | 44.4 (4.8) | 43.4 (4.2) |
| History of CVD, n (%) | 2 (2.3) | 3 (3.1) | 12 (12.6) | 15 (10.1) | 0 (0.0) | 14 (17.9) |
| **Imaging visit** |  |  |  |  |  |  |
| Age (years), median [IQR] | 60.0 [54.0, 66.0] | 57.0 [50.0, 63.0] | 58.0 [53.0, 63.0] | 65.0 [60.8, 69.0] | 57.5 [53.5, 60.0] | 61.5 [57.0, 67.8] |
| Carotid plaque presence, n (%) | 42 (47.7) | 41 (41.8) | 42 (44.2) | 127 (85.8) | 20 (83.3) | 67 (85.9) |
| WMH volume (mL), median [IQR] | 2.4 [1.6, 4.5] | 1.7 [1.2, 2.6] | 1.8 [1.2, 3.3] | 3.6 [2.0, 6.8] | 1.7 [1.0, 3.1] | 2.7 [1.4, 6.0] |
| GM CBF (mL/100g/min), mean (SD) | 57.6 (9.1) | 56.6 (8.3) | 56.0 (9.1) | 55.5 (8.2) | 52.0 (7.8) | 54.4 (8.5) |
| WM CBF (mL/100g/min), mean (SD) | 14.5 (3.2) | 13.5 (3.0) | 12.5 (3.1) | 14.3 (3.4) | 12.1 (3.2) | 13.1 (2.9) |
| GM sCoV (%), mean (SD) | 43.0 (5.7) | 40.6 (4.7) | 43.0 (6.6) | 43.8 (6.3) | 41.5 (5.6) | 42.7 (6.3) |
| WM sCoV (%), mean (SD) | 65.1 (13.8) | 67.7 (14.5) | 76.1 (35.9) | 67.4 (18.9) | 74.3 (26.1) | 71.5 (18.0) |

### Associations between demographic variables and cerebral haemodynamics

Only GM CBF was negatively associated with age (Pearson correlation: -0.15 mL/100g/min per year, 95% CI -0.24; -0.07, P < 0.001, Figure 2). The association between demographics and CBF derived from multivariate regression analyses can be found in Supplementary Table 2A. No associations between sex and GM or WM CBF were observed (P > 0.05). Differences in CBF levels were identified across ethnic groups, with the South-Asian Surinamese having similar GM CBF values compared to their Dutch counterparts, but lower WM CBF values (- 1.57 mL/100g/min, 95% CI -2.26; -0.88, P < 0.001), while individuals of Moroccan descent had lower GM CBF values compared to the Dutch (-1.36 mL/100g/min, 95% CI -2.31; -0.4, P = 0.024).

Only GM sCoV was positively associated with age (0.19% per year, 95% CI 0.11; 0.28, P <0.001, Supplementary Figure 2). Estimates for the association between demographic variables and sCoV as derived from multivariable analyses are available in Supplementary Table 2B. We found significantly lower sCoV values for both WM and GM in females compared to males (-8.34, 95% CI -12.26; -4.41, P < 0.001 for WM and -4.04, 95% CI -5.16; -2.92, P < 0.001 for GM in females). We found slightly higher WM sCoV levels in the South-Asian Surinamese compared to the Dutch (6.89%, 95% CI 2.56; 11.22, P = 0.002).

### Associations between CV risk factors and CBF

For the first CV risk factor visit (Table 2), we found a multivariate association between history of CVD and GM CBF levels (-3.34 mL/100g/min, 95% CI -6.46; -0.22, P = 0.036). Conversely, we found an association between diabetes mellitus and GM CBF in the risk-factor specific models (2.88 mL/100g/min, 95% CI 0.37; 5.39, P = 0.025).

**Table 2:**
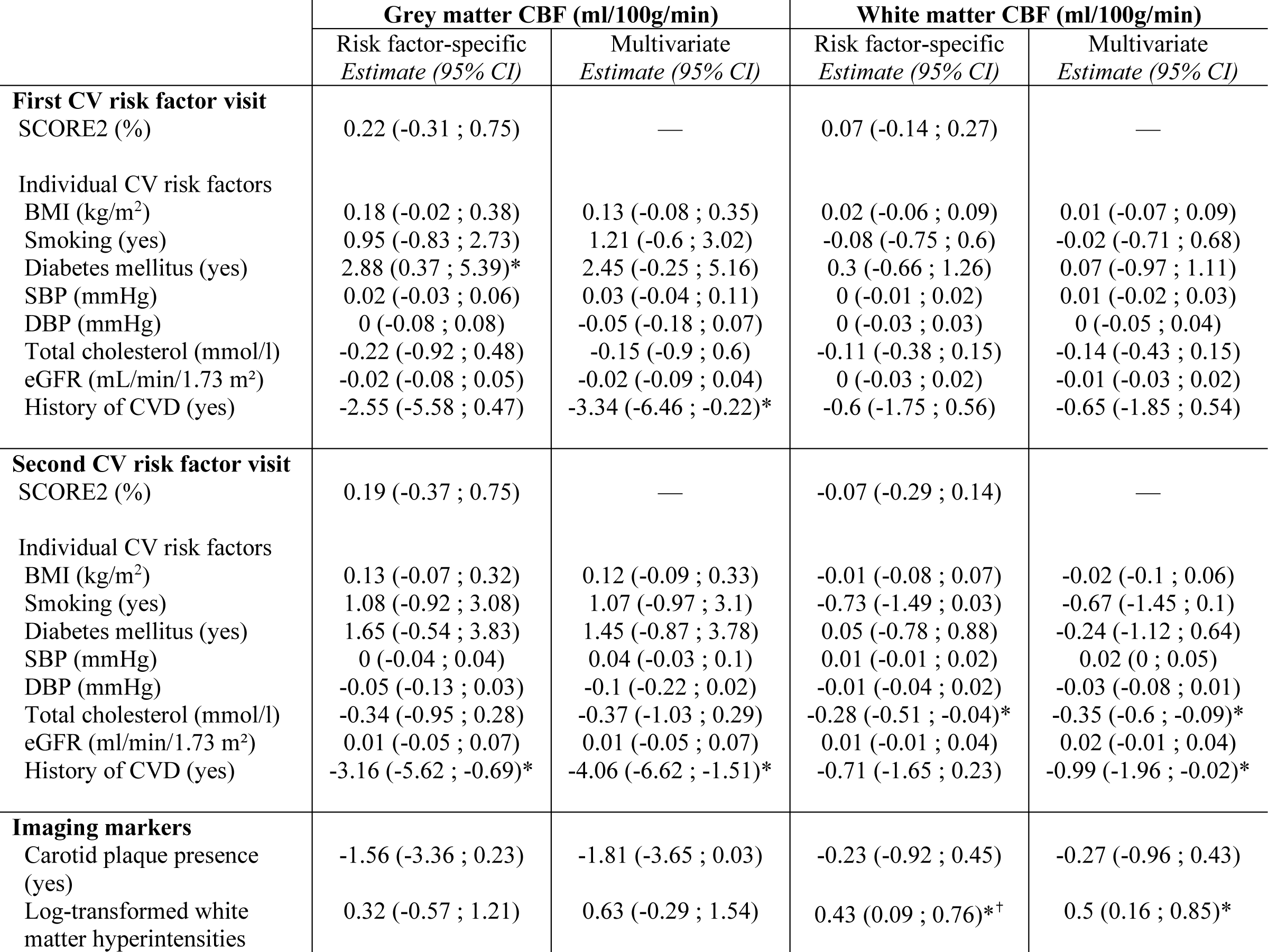
Association of first and second CV risk factor visit with grey and white matter CBF, as derived from linear regression analyses. Risk factor-specific models were adjusted for age, sex, ethnicity, and follow-up time between the CV risk factor visits and MRI measurements. Multivariate models were adjusted for age, sex, ethnicity, follow-up time, BMI, smoking, diabetes mellitus, hypertension, total cholesterol levels, eGFR levels, and a history of CVD. Inverse probability weighting was used to correct for study design. * Statistically significant (P < 0.05). ✝ Statistically significant with FDR correction. BMI = body mass index. CBF = cerebral blood flow. CI = confidence interval. CVD = cardiovascular diseases. DBP = diastolic blood pressure. eGFR = estimated glomerular filtration rate. FDR = false discovery rate. SBP = systolic blood pressure.

|  | Grey matter CBF (ml/100g/min) |  | White matter CBF (ml/100g/min) |  |
| --- | --- | --- | --- | --- |
|  | Risk factor-specific<br>Estimate (95% CI) | Multivariate<br>Estimate (95% CI) | Risk factor-specific<br>Estimate (95% CI) | Multivariate<br>Estimate (95% CI) |
| **First CV risk factor visit** |  |  |  |  |
| SCORE2 (%) | 0.22 (-0.31 ; 0.75) | — | 0.07 (-0.14 ; 0.27) | — |
| Individual CV risk factors |  |  |  |  |
| BMI (kg/m<sup>2</sup>) | 0.18 (-0.02 ; 0.38) | 0.13 (-0.08 ; 0.35) | 0.02 (-0.06 ; 0.09) | 0.01 (-0.07 ; 0.09) |
| Smoking (yes) | 0.95 (-0.83 ; 2.73) | 1.21 (-0.6 ; 3.02) | -0.08 (-0.75 ; 0.6) | -0.02 (-0.71 ; 0.68) |
| Diabetes mellitus (yes) | 2.88 (0.37 ; 5.39)* | 2.45 (-0.25 ; 5.16) | 0.3 (-0.66 ; 1.26) | 0.07 (-0.97 ; 1.11) |
| SBP (mmHg) | 0.02 (-0.03 ; 0.06) | 0.03 (-0.04 ; 0.11) | 0 (-0.01 ; 0.02) | 0.01 (-0.02 ; 0.03) |
| DBP (mmHg) | 0 (-0.08 ; 0.08) | -0.05 (-0.18 ; 0.07) | 0 (-0.03 ; 0.03) | 0 (-0.05 ; 0.04) |
| Total cholesterol (mmol/l) | -0.22 (-0.92 ; 0.48) | -0.15 (-0.9 ; 0.6) | -0.11 (-0.38 ; 0.15) | -0.14 (-0.43 ; 0.15) |
| eGFR (mL/min/1.73 m<sup>2</sup>) | -0.02 (-0.08 ; 0.05) | -0.02 (-0.09 ; 0.04) | 0 (-0.03 ; 0.02) | -0.01 (-0.03 ; 0.02) |
| History of CVD (yes) | -2.55 (-5.58 ; 0.47) | -3.34 (-6.46 ; -0.22)* | -0.6 (-1.75 ; 0.56) | -0.65 (-1.85 ; 0.54) |
| **Second CV risk factor visit** |  |  |  |  |
| SCORE2 (%) | 0.19 (-0.37 ; 0.75) | — | -0.07 (-0.29 ; 0.14) | — |
| Individual CV risk factors |  |  |  |  |
| BMI (kg/m<sup>2</sup>) | 0.13 (-0.07 ; 0.32) | 0.12 (-0.09 ; 0.33) | -0.01 (-0.08 ; 0.07) | -0.02 (-0.1 ; 0.06) |
| Smoking (yes) | 1.08 (-0.92 ; 3.08) | 1.07 (-0.97 ; 3.1) | -0.73 (-1.49 ; 0.03) | -0.67 (-1.45 ; 0.1) |
| Diabetes mellitus (yes) | 1.65 (-0.54 ; 3.83) | 1.45 (-0.87 ; 3.78) | 0.05 (-0.78 ; 0.88) | -0.24 (-1.12 ; 0.64) |
| SBP (mmHg) | 0 (-0.04 ; 0.04) | 0.04 (-0.03 ; 0.1) | 0.01 (-0.01 ; 0.02) | 0.02 (0 ; 0.05) |
| DBP (mmHg) | -0.05 (-0.13 ; 0.03) | -0.1 (-0.22 ; 0.02) | -0.01 (-0.04 ; 0.02) | -0.03 (-0.08 ; 0.01) |
| Total cholesterol (mmol/l) | -0.34 (-0.95 ; 0.28) | -0.37 (-1.03 ; 0.29) | -0.28 (-0.51 ; -0.04)* | -0.35 (-0.6 ; -0.09)* |
| eGFR (ml/min/1.73 m<sup>2</sup>) | 0.01 (-0.05 ; 0.07) | 0.01 (-0.05 ; 0.07) | 0.01 (-0.01 ; 0.04) | 0.02 (-0.01 ; 0.04) |
| History of CVD (yes) | -3.16 (-5.62 ; -0.69)* | -4.06 (-6.62 ; -1.51)* | -0.71 (-1.65 ; 0.23) | -0.99 (-1.96 ; -0.02)* |
| **Imaging markers** |  |  |  |  |
| Carotid plaque presence (yes) | -1.56 (-3.36 ; 0.23) | -1.81 (-3.65 ; 0.03) | -0.23 (-0.92 ; 0.45) | -0.27 (-0.96 ; 0.43) |
| Log-transformed white matter hyperintensities | 0.32 (-0.57 ; 1.21) | 0.63 (-0.29 ; 1.54) | 0.43 (0.09 ; 0.76)*<sup>†</sup> | 0.5 (0.16 ; 0.85)* |

**Table 3:** Association of first and second CV risk factor visit with grey and white matter sCoV, as derived from linear regression analyses. Risk factor-specific models were adjusted for age, sex, ethnicity, and follow-up time between the CV risk factor visits and MRI measurements. Multivariate models were adjusted for age, sex, ethnicity, follow-up time, BMI, smoking, diabetes mellitus, hypertension, total cholesterol levels, eGFR levels, and a positive history of CVD. Inverse probability weighting was used to correct for study design. * Statistically significant (P<0.05). ✝ Statistically significant with FDR correction. BMI = body mass index. CI = confidence interval. CVD = cardiovascular diseases. DBP = diastolic blood pressure. eGFR = estimated glomerular filtration rate. FDR = false discovery rate. SBP = systolic blood pressure. sCoV = spatial coefficient of variation.

|  | Grey matter sCoV (%) |  | White matter sCoV (%) |  |
| --- | --- | --- | --- | --- |
|  | Risk factor-specific<br>*Estimate (95% CI)* | Multivariate<br>*Estimate (95% CI)* | Risk factor-specific<br>*Estimate (95% CI)* | Multivariate<br>*Estimate (95% CI)* |
| **First CV risk factor visit** |  |  |  |  |
| SCORE2 (%) | -0.34 (-0.71 ; 0.03) | — | -0.29 (-1.4 ; 0.82) | — |
| Individual CV risk factors |  |  |  |  |
| BMI (kg/m<sup>2</sup>) | 0.01 (-0.12 ; 0.15) | 0.08 (-0.07 ; 0.23) | 0.29 (-0.19 ; 0.78) | 0.4 (-0.13 ; 0.93) |
| Smoking (yes) | -0.82 (-2.04 ; 0.41) | -0.99 (-2.24 ; 0.26) | 2.1 (-2.19 ; 6.38) | 1.36 (-3.04 ; 5.75) |
| Diabetes mellitus (yes) | -0.76 (-2.5 ; 0.97) | -0.69 (-2.56 ; 1.18) | -0.24 (-6.32 ; 5.83) | -0.62 (-7.2 ; 5.95) |
| SBP (mmHg) | -0.03 (-0.06 ; 0) | -0.07 (-0.12 ; -0.02)* | -0.03 (-0.13 ; 0.08) | -0.13 (-0.31 ; 0.04) |
| DBP (mmHg) | 0 (-0.06 ; 0.05) | 0.09 (0 ; 0.17)* | 0.02 (-0.18 ; 0.21) | 0.16 (-0.14 ; 0.46) |
| Total cholesterol (mmol/l) | -0.07 (-0.56 ; 0.41) | -0.11 (-0.63 ; 0.41) | -0.1 (-1.78 ; 1.59) | 0.25 (-1.57 ; 2.08) |
| eGFR (mL/min/1.73 m<sup>2</sup>) | -0.02 (-0.07 ; 0.02) | -0.02 (-0.06 ; 0.03) | 0.06 (-0.09 ; 0.22) | 0.07 (-0.08 ; 0.23) |
| History of CVD (yes) | 0.16 (-1.94 ; 2.25) | 0.28 (-1.88 ; 2.44) | 6.3 (-1.01 ; 13.6) | 5.5 (-2.08 ; 13.09) |
| **Second CV risk factor visit** |  |  |  |  |
| SCORE2 (%) | -0.12 (-0.51 ; 0.27) | — | 0.95 (-0.45 ; 2.35) | — |
| Individual CV risk factors |  |  |  |  |
| BMI (kg/m<sup>2</sup>) | 0.03 (-0.1 ; 0.17) | 0.09 (-0.05 ; 0.24) | 0.43 (-0.04 ; 0.89) | 0.57 (0.07 ; 1.07)* |
| Smoking (yes) | -0.05 (-1.43 ; 1.34) | -0.08 (-1.47 ; 1.32) | 8.66 (3.89 ; 13.44)*<sup>†</sup> | 8.76 (3.91 ; 13.6)* |
| Diabetes mellitus (yes) | -0.12 (-1.63 ; 1.39) | -0.17 (-1.77 ; 1.43) | 0.12 (-5.16 ; 5.39) | -0.12 (-5.66 ; 5.43) |
| SBP (mmHg) | -0.03 (-0.06 ; 0)* | -0.09 (-0.14 ; -0.05)* | -0.04 (-0.14 ; 0.05) | -0.16 (-0.32 ; -0.01)* |
| DBP (mmHg) | 0.01 (-0.05 ; 0.06) | 0.13 (0.05 ; 0.21)* | 0.01 (-0.17 ; 0.2) | 0.23 (-0.05 ; 0.51) |
| Total cholesterol (mmol/l) | -0.03 (-0.46 ; 0.39) | 0.07 (-0.38 ; 0.52) | 0.75 (-0.72 ; 2.23) | 1.16 (-0.41 ; 2.74) |
| eGFR (ml/min/1.73 m<sup>2</sup>) | -0.04 (-0.08 ; 0.01) | -0.03 (-0.07 ; 0.01) | -0.07 (-0.21 ; 0.07) | -0.07 (-0.21 ; 0.07) |
| History of CVD (yes) | 1.51 (-0.2 ; 3.22) | 1.63 (-0.13 ; 3.39) | 6.45 (0.5 ; 12.4)* | 6.88 (0.78 ; 12.98)* |
| **Imaging markers** |  |  |  |  |
| Carotid plaque presence (yes) | 0.38 (-0.86 ; 1.63) | 0.39 (-0.88 ; 1.65) | 4.39 (0.08 ; 8.71)* | 3.12 (-1.27 ; 7.51) |
| Log-transformed white matter hyperintensities | 0.52 (-0.09 ; 1.13) | 0.55 (-0.08 ; 1.18) | 1.11 (-1.03 ; 3.24) | 0.49 (-1.69 ; 2.68) |

For the second CV risk factor visit (Table 2), we found an association between history of CVD and GM CBF in the risk factor-specific model (-3.16 mL/100g/min, 95% CI -5.62; - 0.69, P = 0.029) and multivariate model (-4.06, 95% CI -6.62; -1.51, P = 0.002). Furthermore, we found an association between total cholesterol levels and WM CBF in both the risk-factor specific (-0.28 mL/100g/min, 95% CI -0.51; -0.04, P = 0.020) and multivariate models (-0.35 mL/100g/min, 95% CI -0.6; -0.09. P = 0.007). No significant associations were found when analysing changes over time in SBP, DBP, and BMI (data not shown).

The observed associations could not be reproduced after stratification for ethnic subgroups (P > 0.05, Supplementary Table 3). Additionally, associations were found of body height with both GM CBF (0.19 mL/100g/min per cm increase in height, 95% CI 0.07; 0.30, p = 0.002), and WM CBF (0.07 mL/100g/min per cm increase in height, 95% CI 0.03; 0.12, p = 0.001).

### Associations between CV risk factors and sCoV

For the first CV risk factor visit, we found an association of SBP levels (-0.07%, 95% CI - 012; -0.02, P = 0.003) and DBP (0.09%, 95% CI 0.0; 0.17, P = 0.040) levels with GM sCoV in the multivariate models only.

For the second CV risk factor visit, SBP levels showed an association with GM sCoV in the risk-factor specific (-0.03%, 95% CI -0.06; 0, P = 0.033) and multivariate models (-0.09%, 95 CI -0.14; -0.05, P < 0.001), whereas DBP levels had an association with GM sCoV only in the multivariate models (0.13%, 95% CI 0.05; 0.21, P = 0.002). Smoking was strongly associated with WM sCoV in both risk-factor specific (8.66%, 95% CI 3.89; 13.44, P < 0.001) and multivariate models (8.76%, 95% CI 3.912; 13.6, P < 0.001). We also found an association between history of CVD and WM sCoV in both the risk-factor specific (6.45%, 95% CI 0.5; 12.4, P = 0.034) and multivariate models (6.88%, 95% CI 0.78; 12.98, P = 0.027) and between SBP levels and WM sCoV in the multivariate models (-0.16%, 95% CI -0.32; - 0.01, P = 0.040) Furthermore, only for change in BMI over time an association was found with WM sCoV (-1.19%, 95% CI -2.26; -0.12, P = 0.030). Lastly, we found an association between body height and GM sCoV (0.08%, 95% CI 0.00; 0.16, P = 0.039).

When exploring the observed associations in the different ethnic subgroups, we found an association of smoking and history of CVD with WM sCoV in the Dutch group, but not in the Moroccan and South-Asian Surinamese subgroup (Supplementary Table 3).

### Associations of carotid plaque and WMH with cerebral haemodynamics

Carotid plaque presence was only associated with WM sCoV in the risk-factor specific model (4.39 mL/100g/min, 95% CI 0.08; 8.71, P = 0.046). WMH was associated with WM CBF in both the risk-factor specific (0.43 mL/100g/min, 95% CI 0.09; 0.76, P = 0.013) and multivariate models (0.5 mL/100g/min, 9% CI -1.96; -0.02, P = 0.004).

### Sensitivity analyses

Substituting the SCORE2 with the Framingham risk score or SBP and DBP with the presence of hypertension, MAP or PP did not reveal associations with CBF. For sCoV, however, it did reveal an association between PP and GM sCoV (data not shown) and an association between hypertension and GM sCoV (data not shown). When adjusting BP values for the effect of anti-hypertensive medication by adding 10 and 5 mmHg to SBP and DBP values in cases where BP-lowering medication was used, SBP remained associated with GM sCoV (data not shown). When adjusting for multiple comparisons, only the associations of WMH volume with WM CBF (P = 0.026) and WM sCoV with smoking (P = 0.003) remained statistically significant in the risk-factor specific models. We found negligible disparities in the associations between CV risk factors and CBF as well as sCoV between models with and without partial volume correction (Supplementary Table 4) and with and without haematocrit correction (Supplementary Table 5). Additionally, the exclusion of participants with a positive history of CVD elicited similar results (Supplementary Table 6).

## Discussion

### Main findings

In this multi-ethnic population-based cohort, we identified several differences in CBF and sCoV between participants of Dutch, Moroccan, and South-Asian Surinamese descent. While we did not find any associations at the first visit, at the second visit, we found mild associations between CVD history and both GM CBF and WM sCoV, total cholesterol levels and WM CBF, SBP and GM sCoV, smoking and WM sCoV, and between WMH volume and WM CBF. No associations with carotid plaque presence were found. Overall, no ethnic differences were found in the association between CV risk factors and CBF or sCoV. Apparently, mild associations start to appear only late in mid-life, are not equally distributed across all risk factors, and appear independent of ethnic CV risk profiles.

### Associations with demographics

Our negative association between age and GM CBF is in agreement with other literature (25, 26), however, our lack of association between age and WM CBF agrees with one study (27) but contrasts with two other studies (28, 29). Differences between these studies might be explained by different age, sex, or ethnicity distributions or by the use of different ROI definitions such as global white matter instead of deep white matter, partial volume effects, ASL acquisitions, or PLD timings (30). Additionally, one study reported slight increases of WM CBF until around 60 years of age (28), suggesting that ageing-related changes in the WM act non-linear in time at older age. This could explain our results, as the median age of our population was 63 years imaging visit.

Consistent with other studies, we found the South-Asian Surinamese sample had a higher prevalence of CV risk factors (31, 32). In addition, they showed a lower WM CBF and higher WM sCoV, implying reduced macrovascular efficiency as a potential consequence of their relatively high CV burden. The fact that we did not observe this in the other participants, suggest that CV risk factors affect cerebral haemodynamics earlier in South-Asian Surinamese. While this might increase the development of WMH (33), we did not observe this in our cohort. Perhaps, the previously reported ethnic disparities in WMH (34-36) occur only after the age of our cohort.

In participants of Moroccan descent, GM CBF was found to be lower than in participants of Dutch descent. Few studies have explored risk factor differences between individuals of Moroccan and European descent (37, 38), some reporting a higher prevalence of CV risk factors in Moroccans (diabetes mellitus) while others indicate lower prevalence (alcohol consumption or smoking). To our knowledge, the present study is the first to assess the relationship between CV risk factors and cerebral haemodynamics in participants of Moroccan descent.

### Associations with CV risk factors

In our study, history of CVD was associated with lower GM CBF in risk factor specific and both lower GM and WM CBF in the multivariate models, which is in line with previous studies correlating low CBF with low cardiac output (39) and stroke (40). Furthermore, a history of CVD was positively associated with WM sCoV, suggesting lower macrovascular efficiency that may have translated to the observed lower WM CBF. These results could imply cerebral haemodynamics are directly affected by (cardio)vascular injury, possibly through changes in cardiac output (41). An alternative indirect explanation to our association is that cardiovascular risk factors equally affect cardiovascular and cerebrovascular circulation. Similarly to one study higher total cholesterol levels were also found to be associated with lower WM CBF (42), which agrees with previous studies correlating LDL cholesterol with WMH load (43) and cardiorespiratory fitness with ATT (44). To what extent such associations are direct or indirect cannot be differentiated with our data.

Our positive association between smoking and WM sCoV is consistent with the finding of a lower WM CBF (45) and decreased localised GM CBF (46, 47) in previous studies. In line with two other studies (15, 48), we found no associations between CBF or sCoV and other risk factors or their composite scores. However, two other studies did report an association between lower GM CBF and higher cardiovascular risk factor composite scores (49, 50). These differences might be attributed to a lower mean age of the population in our study, different ethnic disparities in cardiovascular risk severity present in the aforementioned studies compared to HELIUS, and the use of other ASL MRI parameters.

Higher SBP was related to lower GM sCoV but not GM CBF in our study, in contrast to previous late-life studies that found both hypertension and use of anti-hypertensive medication to be associated with higher sCoV (5, 13) and MAP to be associated with longer arterial transit time (ATT) (40). Perhaps, adequate average perfusion is maintained by improving macrovascular efficiency as a compensatory effect for higher SBP in mid-life.

### Associations with WMH load and carotid plaque

Interestingly, WM CBF showed an unexpected positive association with WMH load. While studies have reported negative associations between WMH load and GM CBF, WM(H) CBF, and WM sCoV (45, 51, 52), positive associations or lack of associations have also been reported (33). The relative low presence of WMH in the mid-life HELIUS cohort compared to other (mostly late-life) studies (14), might explain why a positive association has been found in this study, possibly suggesting a compensatory effect in normal-appearing white matter.

Aside from the low WMH load, indicating an early stage of WMH development, the discrepancies in the findings of our study and others might also be explained by differences in the CBF ROIs. Compared to our deep WM ROI which excluded WMH lesions, other studies may have assessed CBF in total or lobar WM, or within WMH ROIs resulting in different findings (14).

No associations between carotid plaque presence and cerebral haemodynamics were found, contrasting previous studies that show lower CBF (53-55), lower cerebral artery flow (56), more arterial transit artefacts, and increased ATT in patients with carotid stenosis (55, 57). The fact that we could not replicate these associations could be explained by the relatively low prevalence of carotid artery stenosis in mid-life in our relatively healthy population.

### Interactions with ethnicity

We did not find any ethnicity interaction effects on the association between CV risk factors and cerebrovascular haemodynamics, and when stratified for ethnicity, only associations between both smoking and history of CVD with WM sCoV remained for participants of Dutch descent. This fits with the lack of any associations with risk factors at the first CV risk factor visit and the few but relatively weak associations at the second CV risk factor visit. Perhaps, prolonged exposure to CV risk factors affects cerebral haemodynamics only later in life, which seems to be the case for WMH and cognitive decline as well (43, 58, 59). While several (mostly late-life) studies found that associations between CV risk and structural cerebrovascular pathology — in the form of WMH or stroke — differed between ethnicities (60-63), few studies have included cerebral haemodynamics. In one cohort around 62 years of age, ethnicity also did not influence the associations between CV risk factors and perfusion despite ethnic differences in CV risk factor profiles (15). On the other hand, in another cohort around 71 years of age, ethnicity affected the association of whole-brain CBF with memory and executive functioning at 71 years (64). Perhaps, our included study population is too young to detect commonly found cerebral haemodynamic changes from CV risk factors in older populations or any ethnic interaction effects on these associations.

Alternatively, mid-life haemodynamics changes could be too subtle to be picked up with whole-brain resting-state haemodynamics, as one study showed that cumulative FRS between 47 and 67 years was associated with lower GM CBF only in 40% of the GM (50). On the other hand, possible mid-life compensatory effects —such as our positive association between WMH load and WM CBF — might precede and affect the associations that are commonly observed in late-life studies (14). Therefore, future (longitudinal) mid-life studies are suggested to investigate regional cerebral haemodynamics to understand the relationship between CV risk factors, cerebral haemodynamics, and development of WMH, as well as possible compensatory mechanisms.

### Limitations

While we believe our general sample size is sufficient to explore associations between CV risk factors and cerebral haemodynamics, the interaction analysis for detecting ethnic-based disparities in these associations may be underpowered as sample sizes differed per ethnicity. Furthermore, a significant proportion of participants were lost to follow-up. This could result in a non-response bias and overrepresentation of healthier participants in our analyses exploring the effects of CV risk factors on cerebral haemodynamics. Lastly, the use of a deep WM ROI without WMH has the advantage that signal contamination from GM, WMH, and CSF are reduced, associations between WM CBF and WMH volume might behave differently compared to studies that use a less strict WM mask (14).

## Conclusion

In contrast to previous late-life studies, this mid-life multi-ethnic cohort study found several associations between CV risk factors and cerebral haemodynamics at the second visit only. No associations were affected by ethnicity despite differences in their cerebral haemodynamics. Combined with positive associations of CBF and sCoV with WMH volume, these findings indicate that commonly found associations between CV risk factors, cerebral haemodynamics, and WMH might develop only later in life, possibly preceded by compensatory mechanisms. Future studies could extend our work by exploring the link between mid-to-late-life CV risk factors, regional changes in cerebrovascular haemodynamics, and development of WMH in underrepresented populations.

## Author contribution statement

**Esther M.C. Vriend:** Data curation; Investigation; Methodology; Formal Analysis; Visualisation; Writing - Original draft; Writing - Review & Editing.

**Mathijs B.J. Dijsselhof:** Data curation; Investigation; Methodology; Formal Analysis; Visualisation; Writing - Original draft; Writing - Review & Editing.

**Thomas A. Bouwmeester:** Data acquisition; Investigation; Methodology; Conceptualization; Writing - Review & Editing.

**Oscar H. Franco:** Methodology; Conceptualization; Writing - Review & Editing; Funding acquisition.

**Henrike Galenkamp:** Methodology; Conceptualization; Writing - Review & Editing.

**Didier Collard:** Methodology; Conceptualization; Writing - Review & Editing.

**Aart J. Nederveen:** Data curation; Methodology; Writing - Review & Editing.

**Bert-Jan H. van den Born:** Supervision; Writing - Original draft; Writing - Review & Editing; Project administration; Funding acquisition.

**Henk J.M.M. Mutsaerts:** Supervision; Writing - Original draft; Writing - Review & Editing; Project administration; Funding acquisition.

## Supplementary information

Supplementary material for this article is available online.

## Data availability

The HELIUS data are owned by the Amsterdam University Medical Centers, location AMC, in Amsterdam, the Netherlands. Any researcher can request the data by submitting a proposal to the HELIUS Executive Board, as outlined at http://www.heliusstudy.nl/en/researchers/collaboration, by email to. The HELIUS Executive Board will check proposals for compatibility with the general objectives, ethical approvals, and informed consent forms of the HELIUS study. There are no other restrictions to obtaining the data and all data requests will be processed in the same manner. The microbial genomic sequences from the HELIUS cohort, which were used for this study, are stored under protected access on the European Genome-Phenome Archive (https://ega-archive.org/datasets/EGAD00001004106).

## Abbreviations

ASL: arterial spin labelling
ATT: arterial transit time
BMI: body mass index
BP: blood pressure
CBF: cerebral blood flow
cIMT: carotid intima-media thickness
CSF: cerebrospinal fluid
CV: cardiovascular
CVD: cardiovascular disease
DBP: diastolic blood pressure
eGFR: estimated glomerular filtration rate
FA: flip angle
FDR: Benjamini-Hochberg Procedure
FLAIR: fluid-attenuated inversion recovery
FRS: Framingham risk score
GM: grey matter
HDL: high-density lipoprotein cholesterol
HELIUS study: HEalthy Life In an Urban Setting study
LDL: low-density lipoprotein
M0: equilibrium magnetisation image
MAP: mean arterial pressure
MRI: magnetic resonance imaging
PLD: post-labelling delay
PP: pulse pressure
ROIs: regions-of-interests
SBP: systolic blood pressure
SCORE2: Systematic Coronary Risk Evaluation 2
sCoV: spatial coefficient of variation
STROBE: STrengthening the Reporting of OBservational studies in Epidemiology
T1w: T1-weighted
TE: echo time
TI: inversion time
TR: repetition time
WM: white matter
WMH: white matter hyperintensities

